# Characterization of RNA cargo from extracellular vesicles obtained from cerebrospinal fluid and plasma samples in schizophrenia participants and healthy volunteers

**DOI:** 10.1101/2025.01.31.25321299

**Authors:** Juan A. Gallego, Joanna Palade, Eric Alsop, Elizabeth Hutchins, Michael Hsieh, Amber Logerman, Cherae Bilagody, Rebecca Reiman, Bessie Meechoovet, Panieh Terraf, Blake Beecroft, Alex Janss, Francesca Gallaso, Timothy G. Whitsett, Emily A. Blanco, Todd Lencz, Kendall Van Keuren-Jensen, Anil K. Malhotra

## Abstract

Biomarkers that are clinically useful for the diagnosis and treatment of schizophrenia are lacking. Biomarkers are critical tools that reduce the incidence of misdiagnosis, identify subgroups of patients, assist in the proper characterization of patient phenotypes, predict response to treatment or the development of side effects, and can serve as targets for novel therapeutic interventions. In this study, we evaluated small (< 200 nucleotide) and long (> 200 nucleotide) RNAs found in extracellular vesicles (EVs) isolated from the cerebrospinal fluid (CSF) and plasma of individuals with schizophrenia spectrum disorders (SSD) and healthy volunteers (HV). As EVs carry cargo from all tissues in the body, they act as a potential proxy for the tissue of origin, including cells from the brain. We compared the transcriptomic features of EVs from these two biofluids and examined their ability to discriminate between SSD and HV participants, identifying a total of 141 differentially expressed genes, some of which have been previously associated with SSD. Next, we evaluated the potential cell-types that give rise to the SSD-associated CSF RNA cargo, and found the majority were predominantly expressed in excitatory neurons. Our results highlight the potential of EVs as both a source of schizophrenia relevant biomarkers, and molecular insight into disease mechanisms.

## INTRODUCTION

Schizophrenia affects 1% of the population and is the result of complex interplay between genetic and environmental variables, underpinned by dysregulation in neurotransmitter biology, synaptic density and connectivity, and neuroinflammation^1^. The dynamic repertoire of symptoms often coexists with other conditions (Bipolar Disorder, Post-Traumatic Stress Disorder, brain lesions, substance-induced psychosis, etc.), making disease diagnosis and management challenging. Currently, no FDA approved molecular biomarker tests are available for schizophrenia diagnosis^2,3,4,5^.Transcriptomic analysis of human brain tissue has identified significant dysregulation in RNA expression in schizophrenia, offering valuable insight into disease etiology and potential biomarkers^6,7,8,9^. However, molecular work with human samples is generally restricted to postmortem tissue, and the invasive nature of accessing the brain limits the scope of research. While the examination of minimally invasive sources like human blood or peripheral blood mononuclear cells (PBMCs) has found evidence of small RNAs^10,11,12^ and long non-coding transcripts^13,14,15^ associated with schizophrenia, determining if these changes detected in peripheral blood are relevant to a central nervous system disease is challenging. Further, techniques like qRT-PCR and microarrays are limited to predetermined targets, highlighting the need for comprehensive and unbiased methods to explore schizophrenia biomarkers^13,14,15^.

Extracellular vesicles (EVs) are nano-sized, lipid bilayer enclosed particles that shed from all cells and have been detected in all human biofluids. Importantly, EVs contain molecular cargo derived from the cells that released them, and act as a proxy for evaluating changes in affected cells and tissues^16,17^. The EV contents are protected from degradation by the lipid membrane, and may be altered by underlying mechanisms of disease, thus serving as a valuable resource for biomarker discovery. However, the cargo varies depending on the biofluid sampled and the parent tissues that contribute EVs^18,19^. For example, given that the bloodstream receives EVs from many tissues due to the dense vascularization of the body^20^, it can contain proteins and RNAs derived not only from PBMCs and other circulating cells, but also from the brain^21,22,23,24^. Collecting human plasma is minimally invasive, and approximately 30,000 genes are detectable in EVs by RNAseq^25^. Conversely, cerebrospinal fluid (CSF), an ultrafiltrate of plasma generated primarily by cells of the choroid plexus in the brain ventricles, contains only 1-5 cells/mL and is turned over 4-5 times every 24 hours, contributing to lower concentrations of EVs and their cargo in CSF^26^. A key question is whether CSF, despite having fewer detectable genes in EVs (approximately 4,000), might be a superior biofluid for studying schizophrenia biomarkers due to its proximity to the brain.

The majority of EV research on RNAs focuses on small non-coding transcripts: microRNAs (miRNAs), transfer RNA (tRNA), Y RNA fragments, piwi-interacting RNA (piRNA), small nucleolar RNA (snoRNA), and small nuclear RNAs (snRNA); these RNAs are stable and enriched in EVs^27,28^. In addition to small RNAs, there is considerable diversity of long RNAs also present in EVs, both coding and non-coding^29,30^. In this study, we comprehensively examined the small and long RNA contents of plasma and CSF EVs in a cohort of individuals with a schizophrenia-spectrum disorder (SSD) diagnosis and healthy volunteers (HV) using RNA sequencing. While circulating EV cargo has been investigated by other groups in peripheral blood^31,32,33^, to our knowledge, this is the first study to describe CSF EV RNA contents, with participant-matched plasma samples. We hypothesized that small and long RNA transcripts will be differentially expressed in SSD participants compared to HV, and that the repertoire of differentially expressed transcripts in plasma will be different from those in CSF. To understand the relevance of the findings, we conducted Elastic Net modeling, and pathway analysis, in addition to a comparison with brain single nucleus RNA sequencing (snRNA-seq) data.

## METHODS

### Subjects

Individuals with SSD diagnoses were recruited first from the ambulatory outpatient department at The Zucker Hillside Hospital, a Northwell Health stand-alone psychiatric hospital located in Glen Oaks, NY, from 2012 until 2016, and then from the NewYork-Presbyterian Hospital, Westchester Division, in White Plains, NY from May 2016 until May 2019. HV were recruited from the general population via word of mouth, newspaper, internet advertisements, and poster flyers. All subjects provided written informed consent, under a protocol approved by The Institutional Review Boards of the Northwell Health System and the NewYork-Presbyterian Hospital. The PI and the protocol were the same at both institutions since the PI (JAG) moved to the NewYork-Presbyterian Hospital, Westchester Division, in May 2016.

The following requirements were met by all enlisted patients: (1) fulfilled DSM-IV criteria for schizophrenia, schizophreniform disorder, schizoaffective disorder or Psychosis Not Otherwise Specified (NOS); (2) were 15-59 years old, and (3) were willing and capable of providing informed consent. SSD participants were excluded if they (1) were prescribed an anticoagulant, (2) had a history of an organic brain disorder, (3) had a clinically significant thrombocytopenia or coagulopathy based on screening blood tests, or (4) had a diagnosis of a substance-induced psychotic disorder. HV were also excluded if they had (1) an Axis I diagnosis or (2) a first-degree relative with a known or suspected Axis I disorder, as assessed via family history questionnaire. At the time of the participation, none of the participants (either SSD or HV) had any acute medical inflammatory or infectious disease. Psychopathology ratings were administered using the Brief Psychiatric Rating Scale (BPRS), the schedule for assessment of negative symptoms (SANS) and the Clinical Global Impression severity index (CGI-S), and a neuropsychological assessment was administered using the MCCB battery.

### Sample collection and processing

CSF samples were obtained via a lumbar puncture procedure using 25-gauge Whitacre point spinal needles after subcutaneous lidocaine application, using standard techniques, with the participants in a sitting position. All lumbar puncture procedures were conducted between 2 and 3 pm to decrease the potential variability due to circadian rhythm. Fifteen to twenty-five mL of CSF were obtained from each subject, and the first 2-3 ml were used for routine testing (cell count, proteins, and glucose). After collection, CSF samples were centrifuged at 1800 rpm, at 4° for 5 min, to pellet cells. The cell-free supernatant was retrieved, aliquoted, flash frozen with liquid nitrogen, and stored at -80°C. CSF samples with macroscopic blood present were discarded and excluded from this study.

Prior to the lumbar puncture, 10 mL of blood were collected using K2EDTA vacutainer tubes. The blood containers were centrifuged at 2000 rpm, at 4° for 20 minutes, and plasma was retrieved, aliquoted, flash frozen with liquid nitrogen, and stored at -80°C. CSF and blood samples were subsequently sent to the Translational Genomics Research Institute (TGen), in Phoenix, AZ, for EV isolation and analyses.

### EV and RNA isolation

For both CSF and plasma, EV RNA from 1 mL aliquots was isolated using the ExoRNeasy Midi Kit (Qiagen, 77044), according to the manufacturer’s instructions. Aliquots were mixed with binding buffer, applied to the filter column, and washed once. EVs were lysed on-column with QIAzol, followed by phenol-chloroform phase extraction and ethanol precipitation. RNA was applied to the MinElute column, washed with RWT and RPE buffer, and eluted with DNase free RNase free ultrapure H2O. RNA concentration was assessed via Quant-iT RiboGreen (ThermoFisher, R11490).

### Small RNA library preparation and sequencing

Small RNA libraries were generated using NEXTflex Small RNA Library Preparation Kit v2 (Perkin Elmer) from 1 mL of CSF or plasma EV RNA, as previously described^34^. Briefly, samples underwent 3’ adapter ligation, bead purification, 5’ adapter ligation, cDNA synthesis, and a 16 cycle PCR amplification. Finally, size selection was done via gel electrophoresis with 6% polyacrylamide gels, and excision of the 140-160 b.p. nucleotide band, followed by ethanol precipitation. The resulting small RNA libraries were quantified using the High Sensitivity DNA Kit, on an Agilent 2100 Bioanalyzer. Libraries were pooled equimolarly, re-quantified, normalized, denatured at a working concentration range of 6–8 pM with 5% PhiX spike-in, and clustered onto Illumina V3 flow cells using cBot instruments. Sequencing was carried out on Illumina’s HiSeq 2500, for 50 cycles, with TruSeq v3 reagents.

### Whole transcriptome RNA library preparation and sequencing

Up to 3 ng RNA were used for library preparation with SMARTer Stranded Total RNA-Seq Kit v2 - Pico Input Mammalian and SMARTer RNA Unique Dual Index Kits (634411 and 634452, respectively, Takara Bio). RNA was fragmented at 94C for 2 minutes (plasma RNA) or 72C for 3 minutes (CSF RNA), and cDNA was generated with random primers. The first round of PCR was done with 5 cycles, followed by bead purification (AMPure beads, A63882), and ribosomal depletion with targeted ribosome RNA probes. After the second round of PCR with 16 cycles, samples were size selected with beads again, and eluted in ultrapure H2O. The strand-specific, dual-indexed libraries were quantified via qPCR (KAPA SYBR FAST Universal qPCR Kit, KK4824), combined into equimolar pools, and spiked with 1% PhiX. Sequencing was carried out on Illumina’s NovaSeq 6000, with v1.5, 200 cycle kits.

### RNA-sequencing data analysis

For small RNA libraries, the raw sequence image files from the Illumina HiSeq (bcl files) were converted to the fastq format using bcltofastq v. 2.19.1.403 and checked for quality to ensure the quality scores did not deteriorate at the read ends. All samples were processed using the exceRpt small RNA pipeline (PMC7079576) and by-sequence analysis was performed as described previously^35^. Briefly, reads in fastq files were trimmed to remove TruSeq adapters from the 3′ end along with the random 4 bp primers added by the NEXTFLEX v2 kit to the 3′ and 5′ ends using cutadapt. Reads shorter than 15 nucleotides after adapter trimming were discarded. After trimming, the count tables of sequences for each sample were generated by first comparing the adapter trimmed reads against the human genome mapped BAM files from exceRpt to find reads that mapped to the human genome. The genome-mapped reads were then identified in the transcriptome-mapped BAM file to determine the biotypes associated with each read. Since multi-mappers are common with small RNA (short read lengths), biotype was assigned in the priority order used in the exceRpt pipeline: miRNA, YRNA, tRNA, piRNA, and protein-coding biotypes, followed by a bin containing any other annotation in Ensembl (pseudogenes, long non-coding RNAs [lncRNAs], etc.). Reads with the same sequence (after adapter trimming) were then collapsed into a counts table with biotype and gene annotations.

Genes from miRbase (miRNAs), piRNABank (piRNAs), and GtRNAdb (tRNAs) were annotated using the IDs from their respective databases. Protein coding genes and YRNAs from GENCODE are reported as their gene names. All other genes in GENCODE (antisense, long intergenic non-coding RNAs [lincRNAs], misc RNAs, pseudogenes, mitochondrial RNAs, etc.) are reported using their GENCODE transcript IDs. New gene IDs for all gene isoforms were generated by concatenating a number to the gene ID and a look-up table was generated that contains the new gene IDs and the actual gene sequences (Supplementary Table 1). Isoforms were kept as separate transcripts if they differed by one or more nucleotides. We discovered that through our analysis, miR-451a was being incorrectly called asmiR-451b. The correct sequence is reported in the look up tables and the correct mirBase name(miR-451a) was provided throughout the paper.

For long RNA libraries, whole transcriptome data was aligned using STAR v2.6+^36^ to GRCh38 and to additional lncRNAs from the highly curated LNCpedia^37^ and GENCODE datasets. Count data were created using featureCounts^38^.

### Differential Gene Expression Analysis

All count tables (genes counts from whole transcriptome sequencing and sequences from small RNA sequencing) were then loaded into R (v. 4.2.2). Count tables were split into CSF and plasma samples to be analyzed separately. Split counts tables were then filtered to include only genes which were present at >1 count in >50% of samples in the analysis. Differential gene expression analysis was performed in R on the filtered counts tables using DESeq2 (v. 1.38.5) with sex as a covariate, and the standard Benjamini-Hochberg procedure to adjust for multiple comparisons.

### Classification analysis by Elastic Net

Elastic Net (ELNET) is a machine learning tool that leverages aspects of both Lasso and Ridge regression to differentiate between experimental groups, and predict status^39^. We constructed four ELNET models: one for each RNA type (small/WT) in each biofluid (plasma/CSF), using our differentially expressed sequences (p non. adj <0.05). Analyses were performed using the median ratio normalized counts for all sequences from DESeq2, as described previously^35^.

Briefly, following DESeq2 differential expression analysis, sequences with unadjusted *p*-values < 0.05 between sample groups (see above) were selected to be input into the classification model. Modeling was conducted using the glmnet package (version 4.1.7) in R. A series of loops were implemented to (1) identify the optimal lambda value for an elastic net model, (2) select sequences that produced the best performing elastic net model, and (3) estimate performance metrics for the best-performing model. To demonstrate the best model accuracy and generate waterfall plots and heat maps, the best model was tested using all samples and the ability of the model to accurately place each sample was reported using the glmnet predict() function. Additionally, we performed a series of 1000 iterations of glmnet using random 80/20 training/test set splits but kept the lambda and feature set constant (as determined above). This allowed a more robust average accuracy to be calculated using randomized training and testing sets from the reported best model. Waterfall plots and heatmaps containing converters were then generated based on the outcome from the predict() function.

### Pathway analysis

Pathway analysis was performed using the clusterprofiler package in R. Input into cluster profiler was all differentially expressed genes with unadjusted p-value cutoffs set at < 0.05 for CSF and, in order to capture enough significant sequences, < 0.075 for plasma. Pathways reported are from the gene ontology molecular function (GOMF) database. Dot plots were then generated to include all pathways with p-values < 0.05. The top 2 pathways (by lowest p-value) in CSF and plasma analyses were subjected to gene set enrichment analysis (GSEA) using the fgsea R package which produced the barcode plots for these pathways.

### snRNAseq data comparison

Most tissues and cell types secrete EVs that may contribute to the plasma EV milieu^40,41,42,43^. CSF, which bathes the CNS and is separated from the rest of the body by the blood brain barrier (BBB), is likely enriched in EVs originating from the brain and its surrounding structures^44,45,46,47^. We therefore examined whether the DE long RNA genes in CSF EVs could be mapped back to brain cell types using a superior frontal gyrus snRNAseq data set generated from post-mortem control brain samples (n=36, 275,388 nuclei)^48^. Single nuclei data from the superior frontal gyrus was obtained from Synapse^48^ with accession syn51753326. Data was reanalyzed using scanpy as follows: first, data was subset to only controls (n=36) and data integration was rerun using SCVI with 6000 highly variable genes and integrating on sample. Next, UMAP generation and Leiden cluster calling was performed using re-integrated control data. Clusters were then given cell type identities based on well-known marker genes^49^. Expression levels of differentially expressed genes from the control vs SSD in CSF analysis were then explored in the single nuclei data via the generation of violin plots and UMAPs of expression levels to produce a corrected UMAP with cell types labeled using previously published marker genes^49^.

## RESULTS

### Subject Characteristics

Our cohort included 21 SSD participants and 17 HV (n=38 participants). The mean age was 37 years (SD=12.5), and subjects were mostly male (68.4%), which aligns with the increased incidence of schizophrenia in the male population^50^. In the SSD group, 17 subjects (81.0%) had schizophrenia, 3 (14.3%) had schizoaffective disorder and only 1 (4.8%) had psychosis not otherwise specified (Table 1; Supplementary Table 2). Mean BPRS score in the SSD group was 30.5 (SD=10.5). All SSD participants were psychiatrically stable and taking antipsychotic medication at the time of sample collection. There were no statistically significant differences in age, sex, race or ethnicity between SSD and HV groups.

|  | Overall (n=38) | SSD (n=21) | Control (n=17) | p value |
| --- | --- | --- | --- | --- |
| Age, mean (SD) | 37.0 (12.5) | 35.8 (13.9) | 38.4 (10.7) | 0.527 |
| Gender ratio (M/F) | 26/12 | 17/4 | 9/8 | 0.065 |
| Race ratio (Black/White/Other) | 19/13/6 | 12/7/2 | 7/6/4 | 0.769 |
| Diagnosis |  |  |  |  |
| Schizophrenia | n/a | 17 | n/a |  |
| Schizoaffective disorder | n/a | 3 | n/a |  |
| Psychosis NOS | n/a | 1 | n/a |  |
| BPRS test, mean (SD) | n/a | 30.5 (10.5) | n/a |  |
| MCCB composite score, mean (SD) | 37.1 (15.1) | 30.9 (14.4) | 44.8 (12.4) | 0.005 |
| Whole transcriptome RNAseq |  |  |  |  |
| Plasma (n=) | 30 | 16 | 14 |  |
| CSF (n=) | 22 | 12 | 10 |  |
| Small RNAseq |  |  |  |  |
| Plasma (n=) | 35 | 20 | 15 |  |
| CSF (n=) | 34 | 19 | 15 |  |

### Analysis of EV RNA cargo biotypes in plasma and CSF

Small RNAseq was carried out with a ligation-based library generation method that captures the entire small RNA transcript, allowing analyses using all unique sequences, such as isomiRs (a look-up table enumerating each unique gene ID from this study with its corresponding sequence and canonical ID is found in supplementary table 1). We found that the small RNA biotype distribution varied between biofluids. Plasma EVs contained primarily miRNA and YRNA species, whereas tRNA and miRNA comprised 40% and miRNAs 25% of all CSF EVs (Figure 1A). Relative to CSF, plasma small RNAs were more diverse across all biotypes assayed (Figure 1B). Additionally, overlap between the two biofluids was limited to 406 sequences, with the remaining identified 7926 and 325 transcripts exclusive to plasma and CSF, respectively (Figure 1C).

**Figure 1.**
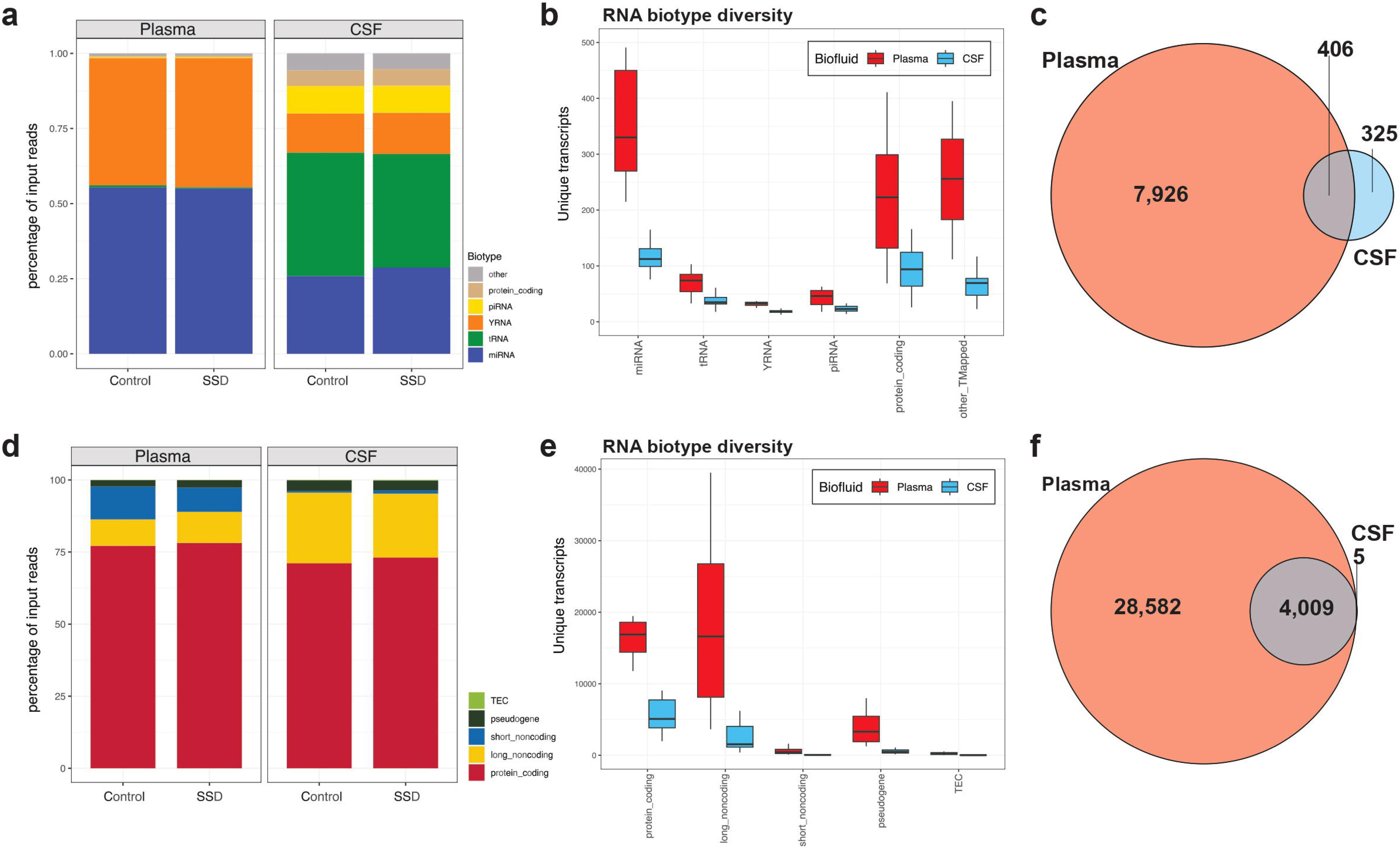
Distribution of small and long RNA cargo in plasma and CSF EVs. The transcriptome mapped reads were each assigned to a biotype category, and displayed as stacked bar plots, separated by disease and control groups, for small RNA (**A**) and long RNA (**D**). Box plots of the total unique genes identified in each RNA biotype showed that plasma EVs (red) were more diverse than CSF EVs (blue) across most biotypes, for small RNA (**B**) and long RNA (**E**). 406 of the 731 CSF small RNAs were shared with plasma small RNA cargo (**C**), but only 5 out of the 4014 long RNA genes identified in CSF EVs were also present in plasma EVs (**F**).

Long RNA was profiled with a workflow that captures coding and noncoding species, maintains strand specificity, and is tailored for low yield sources of RNA. Analysis of long RNA species distribution found that mRNA was the most abundant biotype, making up roughly 75% of all transcriptome-mapped reads in both plasma and CSF (Figure 1D). However, despite the similar biotype distributions, plasma EV RNA had greater overall diversity, with few unique genes identified exclusively in the CSF EVs (n=5) (Figures 1E and 1F). Finally, PCA examining how different attributes (sex, age, race) drive variability in gene expression for both small and long RNA found no significant associations, except for sex, in the long RNA plasma dataset (Supplementary Figure 1). Of note, sample/patient IDs present in Supplementary Figures 1 & 2 were not known to anyone outside the research group. For subsequent differential expression (DE) analysis of the long RNAs, sex was used as a covariate.

### Differentially expressed transcripts in plasma and CSF EVs

Differential expression analysis of small RNA transcripts in plasma showed that out of 8,332 sequences detected, 81 were statistically significantly different (p adj. <0.05), the majority belonging to the miRNA (79%) and YRNA (10%) biotypes (Figure 2A; Supplementary File 3). Notably, 9 isoforms of miR-451a were significantly upregulated in SSD, while sequences of miR-223 and miR-146a were downregulated in our disease group (Figure 2B). Analyses of small RNA fragments in CSF EVs showed that 12 sequences were significantly different at p adj.<0.05), with 11 of them downregulated in SSD, and one upregulated (miR-181a) (Figure 2D; Supplementary File 3).

**Figure 2.**
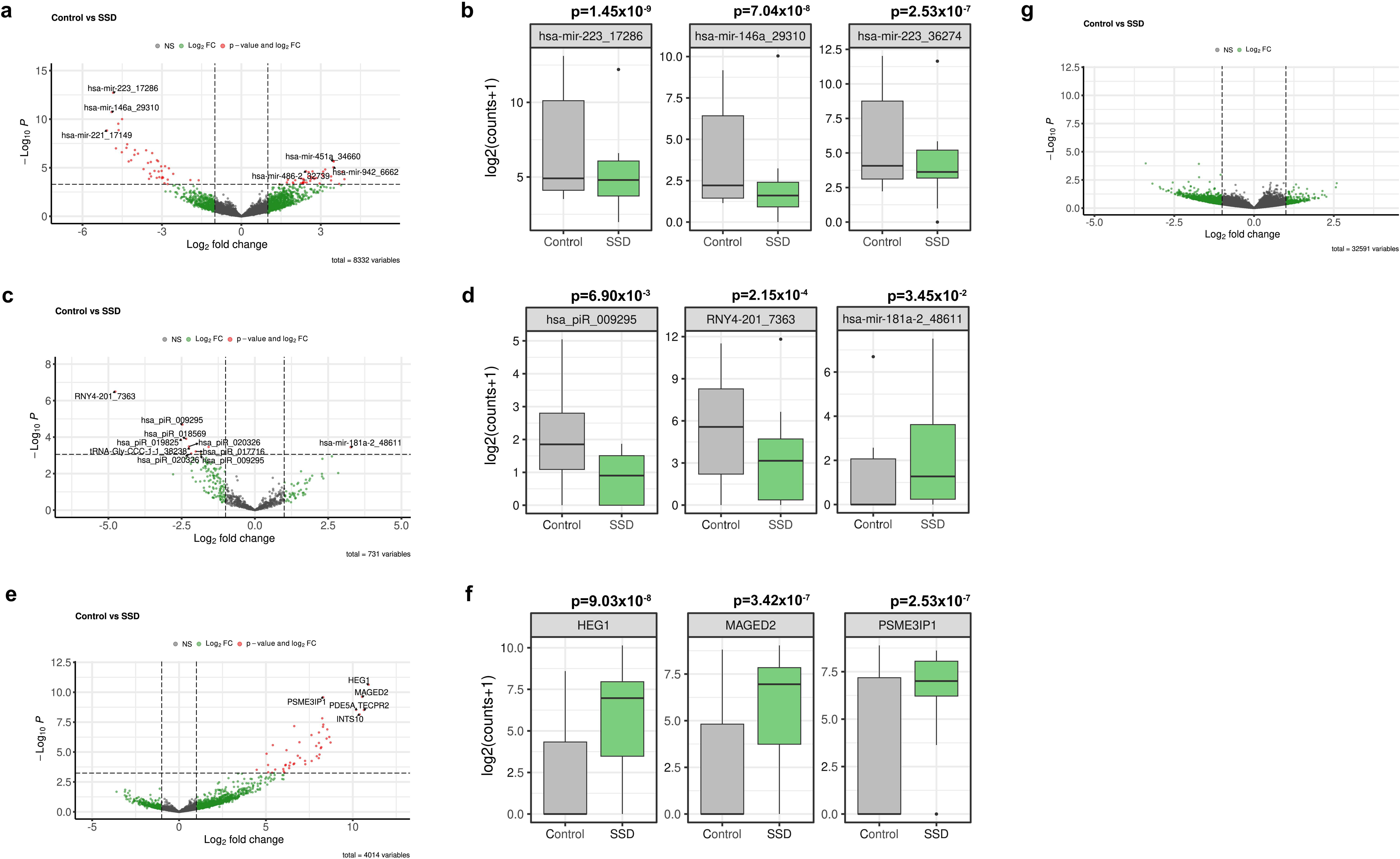
Differentially expressed RNA plasma and CSF EV cargo in SSD. Differential gene expression of the small RNA repertoire found 81 significant transcripts in plasma EVs **(A)**, and 12 significant transcripts in CSF EVs **(D)**. Analysis of long RNA found 48 significant genes in CSF EV, all upregulated in SSD. Box plots show the top 3 most significantly downregulated and upregulated genes in each comparison with corresponding p adj. values, for small RNA plasma EVs **(B, C)**, small RNA CSF EVs **(E)** and long RNA CSF EVs **(G)**. ELNET model of participants with both small and long RNAseq (12 control and 15 SSD samples) data used 18 genes to differentiate between groups **(H)**. The waterfall plot across the top of the heatmap **(H)** demonstrates ELNET model confidence in assigning groups (0 to 1, with 0.5 being no confidence in assignment), with dots colored to the diagnosis group (gray=control, green=SSD).

Whole transcriptome analysis of CSF EVs found 48 differentially expressed genes at p adj <0.05, all of them were upregulated in SSD (Figure 2F, Supplementary File 3). Most of the significant sequences were protein coding, such as *PSME3IP1*, which encodes a proteasomal complex component (Figure 2G). In plasma, DE analysis of long RNA did not yield significant transcripts when adjusting for multiple comparisons (Supplementary file 3).

### Elastic Net Analysis

In the CSF/small ELNET analysis, the model chose 14 transcripts, and of those 8 were miRNAs (miR-7b, miR-7c, miR10a, miR-26b, miR-92a-1, miR-152, miR-204, miR-486-2) with an Area Under the Curve (AUC) of 0.78 (SD=0.02). In the plasma/small ELNET model, 12 transcripts were selected (AUC: 0.73, SD=0.04) and of those 5 were miRNAs (miR-7g, miR-93, miR-128, miR-374c and miR-574). In the CSF/WT model, 11 transcripts were selected with an AUC=0.76 (SD=0.21) and in the plasma/WT model, 12 transcripts were selected (AUC: 0.87, SD=0.02). (Supplementary Figure 2).

### Pathway analysis of DE genes in CSF and plasma EVs

Examining the differentially expressed EV RNA sequences in our cohort hinted at potential SSD dysregulation in various neuronal processes, immune regulation, and proteostasis. Integrated pathway analysis showed that cytokine signaling was significant in plasma EVs, with the corresponding genes downregulated in SSD (Figure 3A and 3B), whereas genes associated with nicotinamide adenine dinucleotide (NAD) binding were upregulated in CSF (Figure 3C and 3D). In both biofluids, protein ubiquitination processes were significantly enriched, particularly in CSF, with the repertoire of pathway associated genes significantly upregulated in SSD.

**Figure 3.**
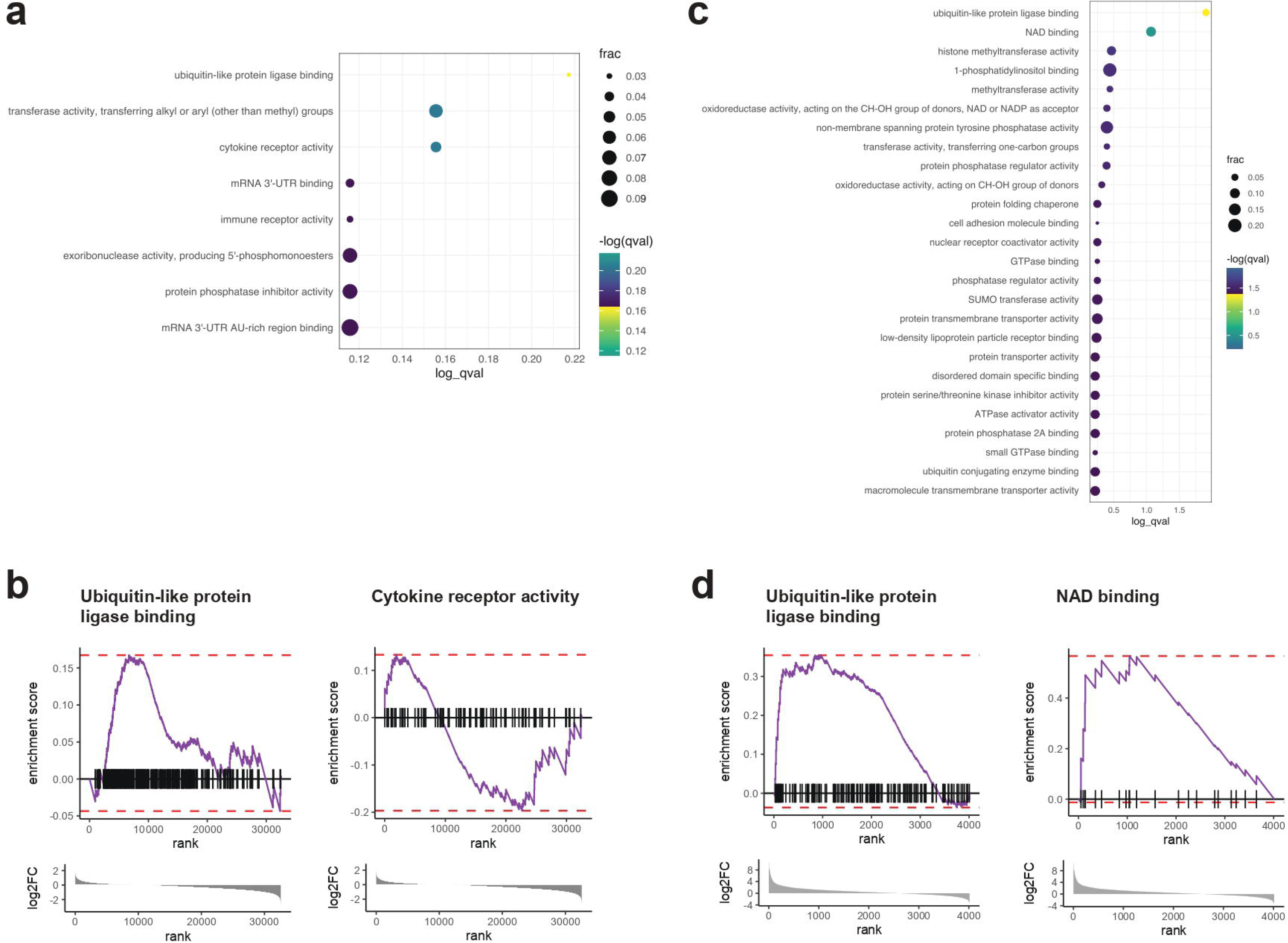
Pathway analysis of differentially expressed long RNA transcripts in plasma and CSF EVs. Pathway analysis in plasma **(A)** and CSF **(C)** EVs for “molecular function” was done with DE transcripts (p<0.075 for plasma, p<0.05 for CSF, non-adjusted), and shown as dot plots. Expression of genes in individual enriched pathways was visualized in barcode plots for plasma **(B)** and CSF **(D)** pathways. In the dot plots, dot size and color indicate the number of genes and the statistical significance, respectively.

### snRNAseq data comparison of differentially detected CSF genes

All 48 long RNA differentially expressed genes in whole transcriptome RNA CSF analyses were identified in the snRNAseq data, and 32 were expressed robustly enough to reach a median expression greater than zero in at least one cell cluster. Strikingly, 75% of the transcripts had the highest expression in the excitatory neuron cluster, with 15 of them exclusively reaching a median expression > zero in excitatory neurons (Table 2; Figure 4A and 4B). Expression of 4 genes was highest in the microglia cluster, including *IL6ST*, which encodes cytokine receptor CD130 (Table 2; Figure 4D). Together, these findings suggest that the CSF EV whole transcriptome RNA signature in SSD is primarily associated with RNAs from excitatory neurons in the frontal cortex, and to a lesser extent, microglia.

**Figure 4.**
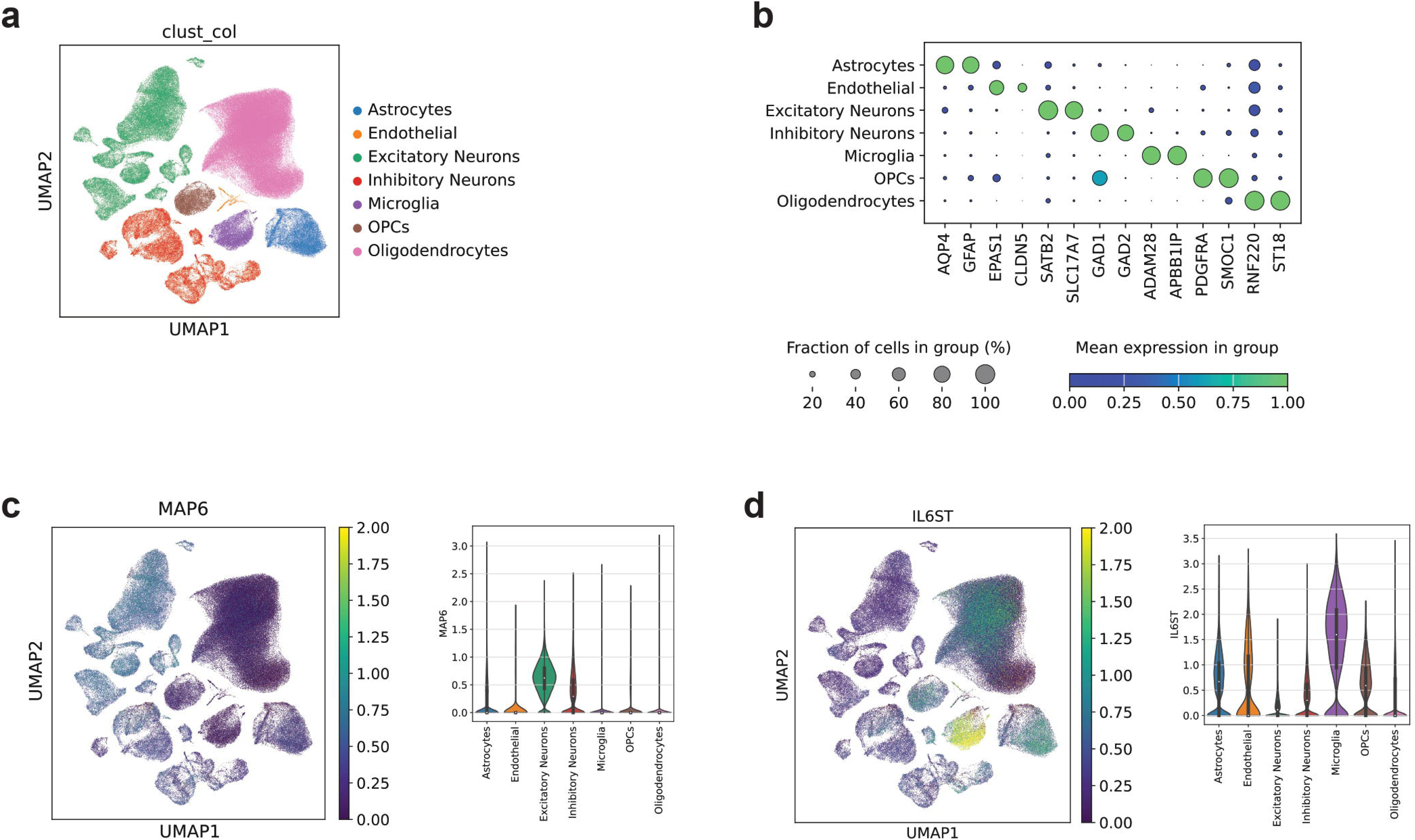
CSF EV long RNA transcripts expression in brain cell types. snRNAseq data from 36 control participants and a total of 275,388 nuclei are represented in a UMAP graph **(A)**. Cell identity per each cluster was assigned based on expression of canonical marker genes, as shown in a dot plot **(B)**. CSF EV transcripts specific to excitatory neurons **(C)** and upregulated in microglia **(D)** are shown as violin plots and UMAPs. On the violin plots, the Y axis denotes the normalized counts per cell.

**Table 2.**
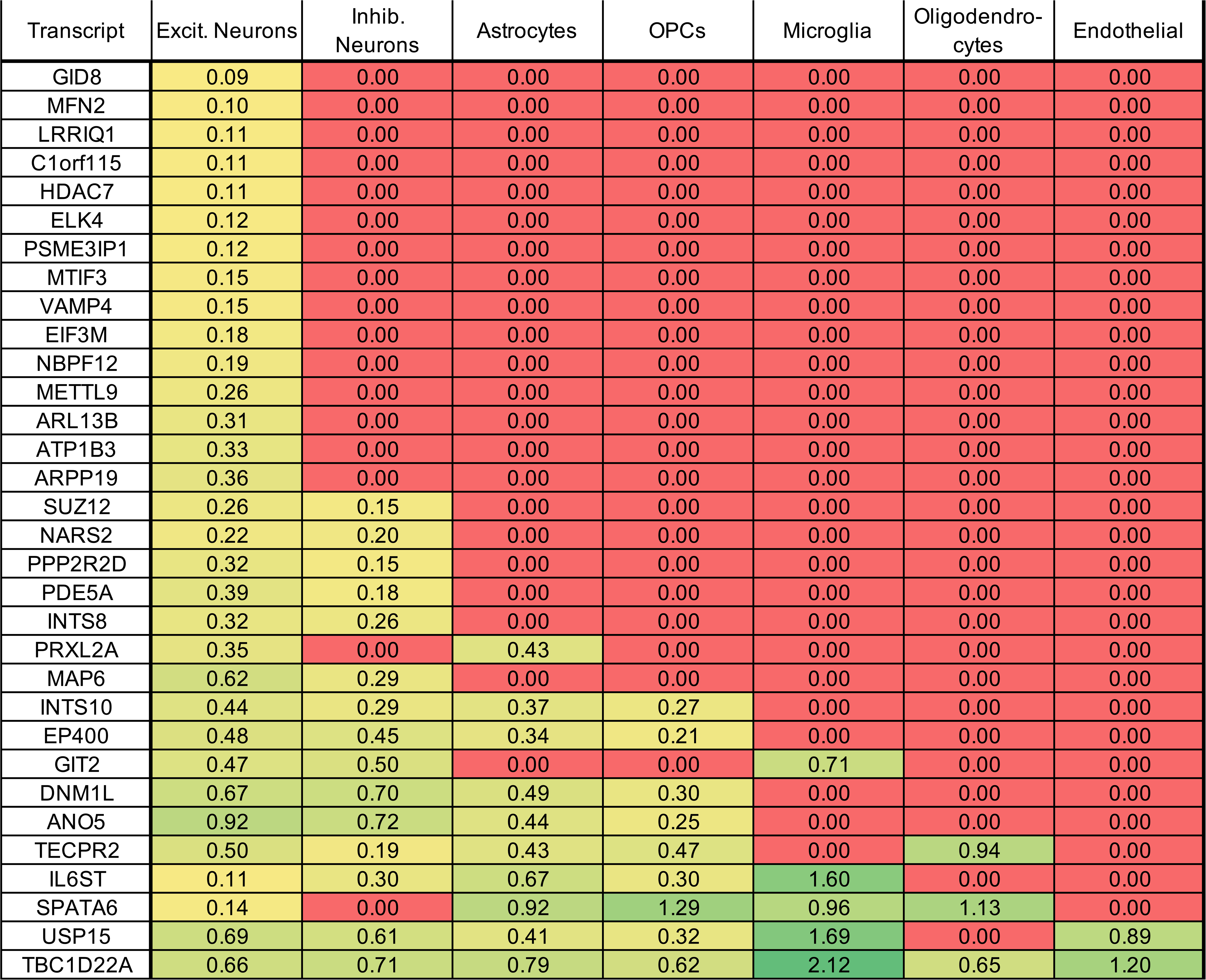
DE CSF EV transcripts in the brain. The significantly DE genes from the Control vs. SSD analysis were examined in the snRNAseq human prefrontal cortex data set. For each transcript, the median expression level in each cell cluster is presented with cells colored on a red-green scale proportional to the median depicted. Transcripts that did not reach median >0 in any cell cluster were not included.

| Transcript | Excit. Neurons | Inhib. Neurons | Astrocytes | OPCs | Microglia | Oligodendro-cytes | Endothelial |
| --- | --- | --- | --- | --- | --- | --- | --- |
| GID8 | 0.09 | 0.00 | 0.00 | 0.00 | 0.00 | 0.00 | 0.00 |
| MFN2 | 0.10 | 0.00 | 0.00 | 0.00 | 0.00 | 0.00 | 0.00 |
| LRRIQ1 | 0.11 | 0.00 | 0.00 | 0.00 | 0.00 | 0.00 | 0.00 |
| C1orf115 | 0.11 | 0.00 | 0.00 | 0.00 | 0.00 | 0.00 | 0.00 |
| HDAC7 | 0.11 | 0.00 | 0.00 | 0.00 | 0.00 | 0.00 | 0.00 |
| ELK4 | 0.12 | 0.00 | 0.00 | 0.00 | 0.00 | 0.00 | 0.00 |
| PSME3IP1 | 0.12 | 0.00 | 0.00 | 0.00 | 0.00 | 0.00 | 0.00 |
| MTIF3 | 0.15 | 0.00 | 0.00 | 0.00 | 0.00 | 0.00 | 0.00 |
| VAMP4 | 0.15 | 0.00 | 0.00 | 0.00 | 0.00 | 0.00 | 0.00 |
| EIF3M | 0.18 | 0.00 | 0.00 | 0.00 | 0.00 | 0.00 | 0.00 |
| NBPF12 | 0.19 | 0.00 | 0.00 | 0.00 | 0.00 | 0.00 | 0.00 |
| METTL9 | 0.26 | 0.00 | 0.00 | 0.00 | 0.00 | 0.00 | 0.00 |
| ARL13B | 0.31 | 0.00 | 0.00 | 0.00 | 0.00 | 0.00 | 0.00 |
| ATP1B3 | 0.33 | 0.00 | 0.00 | 0.00 | 0.00 | 0.00 | 0.00 |
| ARPP19 | 0.36 | 0.00 | 0.00 | 0.00 | 0.00 | 0.00 | 0.00 |
| SUZ12 | 0.26 | 0.15 | 0.00 | 0.00 | 0.00 | 0.00 | 0.00 |
| NARS2 | 0.22 | 0.20 | 0.00 | 0.00 | 0.00 | 0.00 | 0.00 |
| PPP2R2D | 0.32 | 0.15 | 0.00 | 0.00 | 0.00 | 0.00 | 0.00 |
| PDE5A | 0.39 | 0.18 | 0.00 | 0.00 | 0.00 | 0.00 | 0.00 |
| INTS8 | 0.32 | 0.26 | 0.00 | 0.00 | 0.00 | 0.00 | 0.00 |
| PRXL2A | 0.35 | 0.00 | 0.43 | 0.00 | 0.00 | 0.00 | 0.00 |
| MAP6 | 0.62 | 0.29 | 0.00 | 0.00 | 0.00 | 0.00 | 0.00 |
| INTS10 | 0.44 | 0.29 | 0.37 | 0.27 | 0.00 | 0.00 | 0.00 |
| EP400 | 0.48 | 0.45 | 0.34 | 0.21 | 0.00 | 0.00 | 0.00 |
| GIT2 | 0.47 | 0.50 | 0.00 | 0.00 | 0.71 | 0.00 | 0.00 |
| DNM1L | 0.67 | 0.70 | 0.49 | 0.30 | 0.00 | 0.00 | 0.00 |
| ANO5 | 0.92 | 0.72 | 0.44 | 0.25 | 0.00 | 0.00 | 0.00 |
| TECPR2 | 0.50 | 0.19 | 0.43 | 0.47 | 0.00 | 0.94 | 0.00 |
| IL6ST | 0.11 | 0.30 | 0.67 | 0.30 | 1.60 | 0.00 | 0.00 |
| SPATA6 | 0.14 | 0.00 | 0.92 | 1.29 | 0.96 | 1.13 | 0.00 |
| USP15 | 0.69 | 0.61 | 0.41 | 0.32 | 1.69 | 0.00 | 0.89 |
| TBC1D22A | 0.66 | 0.71 | 0.79 | 0.62 | 2.12 | 0.65 | 1.20 |

## DISCUSSION

Sampling human biofluids and interrogating their contents could be tremendously useful for psychiatric conditions, offering insight into molecular mechanisms of disease and biomarker development. The EV fraction of human biofluids is of particular interest, as it may provide a snapshot of the inner workings of tissues like the CNS that are otherwise difficult to access. Indeed, numerous studies have examined human biofluid EVs for cargo associated with neurodegenerative conditions (Alzheimer’s disease, Parkinson’s disease), although research into their relevance for schizophrenia remains scarce^51,52,53,54,33^. In this study, we used RNAseq to describe the full repertoire of EV RNA cargo in human plasma and CSF, with library preparation workflows uniquely tailored for cell-free samples, in a cohort of 21 SSD patients and 17 HV.

Alterations in miRNA biology in SSD have been reported, as evidenced by a GWAS finding disease-associated SNPs in mir-137^55^ or miRNA processing machinery^56^. Other studies have found dysregulated miRNA expression in post-mortem brains^57,58,59^ or peripheral blood^60,61,62^. In our cohort, the small RNA content distribution in EVs varied depending on the biofluid sampled, as we described previously^63^, with miRNA and YRNAs enriched in plasma EVs, while miRNA and tRNAs (Figure 1) transcripts were abundant in CSF EVs (Figure 1A-C). DE analysis between cases and controls found transcripts that were both novel, and previously associated with neuropsychiatric disorders (Figure 2). In CSF, 8 DE small RNA fragments were piRNAs, 2 were tRNA and 1 was a miRNA. Out of those transcripts, only miR-181a has been previously associated with schizophrenia, in a study that used a bioinformatics approach with data obtained from the Gene Expression Omnibus database to identify miRNAs targeting lncRNAs extracted from post-mortem brain tissues of the BA10 area from SCZ and healthy participants, and not by assessing CSF^64^.

In plasma, out of 81 DE small RNA sequences, 65 were miRNAs, 7 were YRNAs, 2 were tRNAs, 2 were piRNAs, and the rest were miscellaneous mapped sequences. Out of the 65 DE miRNAs, several have been previously associated with schizophrenia. Ibrahim et al reported an association between a miR-146a C > G single nucleotide polymorphism (SNP rs2910164) and schizophrenia individuals with and without an acute ischemic stroke compared to healthy controls^65^. miR-143 and miR-486 were reported by Tomita and colleagues, who used urinary exosomal miRNAs to predict persistent psychotic-like experiences in 345 teens^66^.

Similarly, Sargazy and colleagues^67^ reported an SNP (rs4705343T/C) in the promoter region of mir-143 associated with an increased risk of schizophrenia. Sabaie et al found that <u>miR-26a</u> was also one of the seven miRNAs included in lncRNA-miRNA-mRNAs axes^64^. Beveridge and colleagues found upregulation of miR-15b, miR16 and <u>miR-26b</u> in human superior temporal gyrus (STG) tissues^58^, (all of them significant in our cohort)^58^. In another paper by Sabaie and colleagues^68^ they used a bioinformatics approach to describe the role of long non-coding RNA (lncRNA)-associated competing endogenous RNAs (ceRNAs) in the olfactory epithelium (OE) biopsy samples from schizophrenia individuals and healthy controls and found six potential DElncRNA-miRNA-DEmRNA loops involved in the pathophysiology of schizophrenia. One of those loops comprised the *SOX2* overlapping transcript, <u>miR-24-3p</u> and *NOS3*. Lastly, in a study by Feng and colleagues^69^, also using bioinformatic analysis to create gene-miRNA interaction networks, they studied the correlation between ferroptosis (i.e., a type of programmed cell death that includes accumulation of iron and lipid peroxides) genes and schizophrenia and found that <u>miR-16-5p</u> was related to three hub genes: tumor protein p53 (*TP53*), Vascular endothelial growth factor A (*VEGFA*) and Prostaglandin-endoperoxide synthase 2 (*PTGS2*). Importantly, in a very recent meta-analysis of miRNAs in peripheral blood in schizophrenia^12^, the authors arrived at 10 candidate schizophrenia-associated miRNAs by applying two computational learning methods, 4 of which were DE in our study (<u>miR-16</u>, <u>miR-146a</u>, <u>miR-221</u> and <u>miR-222</u>). Numerous piRNAs and YRNA transcripts were DE in CSF and plasma in our cohort. However, research into the biological significance of these RNA biotypes is still emerging, making their relevance to disease unclear.

In our whole transcriptome analyses, we did not find any DE genes between SSD and HV in plasma, but we detected 48 DE genes in CSF. The EV long RNA landscape was dominated by protein coding genes, regardless of disease status (Figure 1A-C). Most significantly different genes were upregulated in disease, and many were associated with post translational modification, ubiquitination and degradation. About a quarter of the DE genes detected in our study have been previously associated with schizophrenia. IL-6 signal transducer (*IL6ST*) was found to be increased in midbrain of human postmortem brain tissue from 28 SZ individuals and 29 healthy controls^70^. Mitofusin 2 (*MFN2*) a molecule responsible for mitochondrial morphology and metabolism, was found in lower quantities in astrocyte-derived extracellular vesicles and neuron-derived extracellular vesicles obtained from plasma in 10 first episode psychosis individuals compared to 10 healthy controls. Similarly, a blood based methylome-wide association study identified altered methylation at the *MFN2* locus in patients with schizophrenia^71^. However, protein levels of MFN2 assayed using western blot were not significantly different in post-mortem rostral anterior cingulate gyrus tissue in a cohort of 25 SZ and 13 normal controls^72^ and *MFN2* mRNA was downregulated in PBMCs from 60 SZ participants compared to 20 healthy controls, and in lymphoblastoid cell lines treated with clozapine or olanzapine^73^. Two SNPs in Microtubule-associated protein 6 (*MAP6*), which was upregulated in CSF EVs in our disease cohort, were significantly associated with schizophrenia, and of the two MAP isoforms measured in human post-mortem prefrontal cortex (BA 46), one was significantly upregulated in SZ patients^74^. On the other hand, Shao and colleagues measured MAP6 protein levels using enzyme-linked immunosorbent assay (ELISA) in subcortical white matter from the DLPFC (BA 9) from 35 SZ individuals, 34 bipolar disorder and 35 healthy controls^75^ and found significant lower levels in bipolar disorder individuals compared to healthy controls, with levels also lower in SZ individuals without reaching statistical significance. The Chromosome 1 Open Reading Frame 15 (*C1orf115*) gene was found to fulfill Mendelian Randomization criteria for putative causality for psychosis biotypes, along with other 3 genes and transcripts as part of a study that conducted a genomic analysis of biotypes as part of the Bipolar-Schizophrenia Network for Intermediate Phenotypes (B-SNIP)^76^. In a study comparing expression levels of 12 different genes across clinical high risk (CHR), first episode psychosis (FEP), chronic schizophrenia (CSZ) and healthy controls (HC), the Ubiquitin Fusion Degradation 1 (*UFD1*) was upregulated in CSZ compared to the HCs and FEP and in CHR compared to HCs and FEP^77^. Moreover, SNPs in the *UFD1* gene have been consistently associated with schizophrenia^78,79,80^. In terms of the TBC1 Domain Family Member 22A (*TBC1D22A*) gene, CpG sites in this gene were hypomethylated in SZ participants compared to healthy controls^81^. *DNM1L* (a.k.a drp-1) was downregulated in PBMC from 60 patients with SZ and 20 HC and was also found to undergo olanzapine and clozapine induced downregulation in lymphoblastoid cell lines^73^. *PSMA4* was proposed as a causal gene for schizophrenia along with other six genes in a study by Ma and colleagues, where they integrated the results from different prediction approaches. Similarly, genetic variants on 15q25 that were associated with higher smoking quantity and schizophrenia were also associated with *PSMA4* expression in specific brain regions^82^. In addition, *PSMA4* was found to interact with dysbindin (*DTNBP1*), a gene frequently associated with schizophrenia^83^ and was associated with smoking behavior and schizophrenia using local genetic correlation analysis^84^. *SUZ12*, a transcription factor and another DE transcript in our study, was selected using transcription factor enrichment analysis as one of the top transcription factors regulating DE genes in SZ^85^. Lastly, *GIT2* was found to be one of 10 hypermethylated genes in a schizophrenia differential methylation network that might be important in the regulation of gene expression in schizophrenia^86^.

Despite our limited sample size, ELNET models provided moderate diagnostic accuracy with AUCs that ranged between 0.73 and 0.87, depending on which RNA type and fluid was investigated. Interestingly, several miRNAs that have been associated with schizophrenia were selected by these models, such as miR-26b^87,88^, ( miR-92a^89,90,91,92^, miR-204^93^ and miR-486^94,90^. Although a validation of these models in a much larger group of individuals, and ideally in first episode individuals, would be required, these data provide a strong starting point in our goal to find a feasible, clinically useful and scalable diagnostic biomarker for schizophrenia.

Research into molecular mechanisms of disease has outlined substantial involvement of dopaminergic^95^ and glutamatergic neurons^96^ in SSD, as well as altered astrocyte function and microglial activation^97^. While affected brain cells might be the source of the DE genes associated with disease, it is difficult to pinpoint the origins of EVs from peripheral circulation, which perfuses most internal structures, although attempts to investigate brain-derived EVs have been made^98,99^. CSF contents are restricted by the BBB and likely to be enriched in CNS-specific EVs. We explored the origin of the most significantly DE EV cargo in CSF using a human brain snRNAseq data set (Figure 5). Out of the 32 genes that reached a median cell expression >0 in at least one cluster, 24 were most highly expressed in excitatory neurons, with some genes (like *METTL9* or *ARPP19*) exclusively reaching a median greater than zero in the excitatory cluster. Microglia were also represented, with 4 out of 32 genes (including *IL6ST* and *USP15*). These findings suggest the DE transcripts in CSF EVs from SSD could be predominantly neuronally derived and are consistent with our current understanding of disease etiology. Furthermore, they align with recent, large-scale snRNAseq analyses of schizophrenia and control post-mortem brains where the majority of DE genes are found in excitatory neuron clusters^100^.

Our study is the first to use sequencing to examine the full repertoire of small and long, coding and noncoding RNA in matched human plasma and CSF EVs in schizophrenia, and map disease associated genes back to specific CNS cell types using brain snRNAseq data. RNAseq allows us to examine the full EV RNA landscape without relying on pre-determined targets, generate unbiased diagnosis models, and identify novel disease associated transcripts.

However, we recognize several limitations in our study, such as the limited number of participants (n=38). Also, our SSD patients were on antipsychotic medication at the time of sample collection, and it is not possible to discern which transcripts were influenced by disease, treatment, or both. Finally, it is also possible that some of our findings are relevant to other psychiatric disorders, such as bipolar disorder (BPD) or major depressive disorder (MDD) since it is demonstrated that they share genetic and biological factors with SSDs^101,102^. Our findings should be validated in a larger cohort, ideally featuring longitudinal samples taken from various time points and conditions, including antipsychotic-naïve, first episode psychosis cases, and compared against patients with BPD and MDD. Together, our data suggests that quantifiable, significantly DE transcripts related to immune dysregulation and neuronal function are present in biofluid EVs, particularly in CSF. These changes could be leveraged for biomarker development, to diagnose and monitor psychiatric conditions.

## Data Availability

All data produced in the present study are available upon reasonable request to the authors.

## Acknowledgements

The authors wish to acknowledge Christopher Morell for helping with data collection, Santiago Vega for helping with manuscript formatting and Jeff Watkins for figure formatting and editing in Adobe Illustrator.

## Figure legends

**Supplemental Figure 1. Distribution of long and small RNA cargo in plasma and CSF EVs in each sample.** The mapping summary percentages for small RNA **(A)** and overall biotype distribution **(C)** were shown for each participant, and each biofluid. The percentage of long RNA input reads for each sample mapping to the transcriptome **(B)** and the biotype distribution within each set of transcriptome-mapped reads **(D)** were shown as stacked bar plots. Principal component analysis for small RNA **(E)** and long RNA **(F)** found sex as a significant variable for plasma EV long RNA in PC3 (p< 0.001). TEC= to be experimentally confirmed; UniVec= library adapter reads; ethnic= Hispanic or Latino Y/N.

**Figure 2 supplemental. CSF EV long RNA transcripts expression in brain cell types.** Additional examples of transcripts from the CSF EV signature mapped onto the brain snRNAseq data set are shown as violin plots and UMAPs. On the violin plots, the Y axis denotes the normalized counts per cell.

