## Supplementary figures and images for "Characterization of RNA cargo from extracellular vesicles obtained from cerebrospinal fluid and plasma samples in schizophrenia participants and healthy volunteers"

### Supplementary Figure 1.

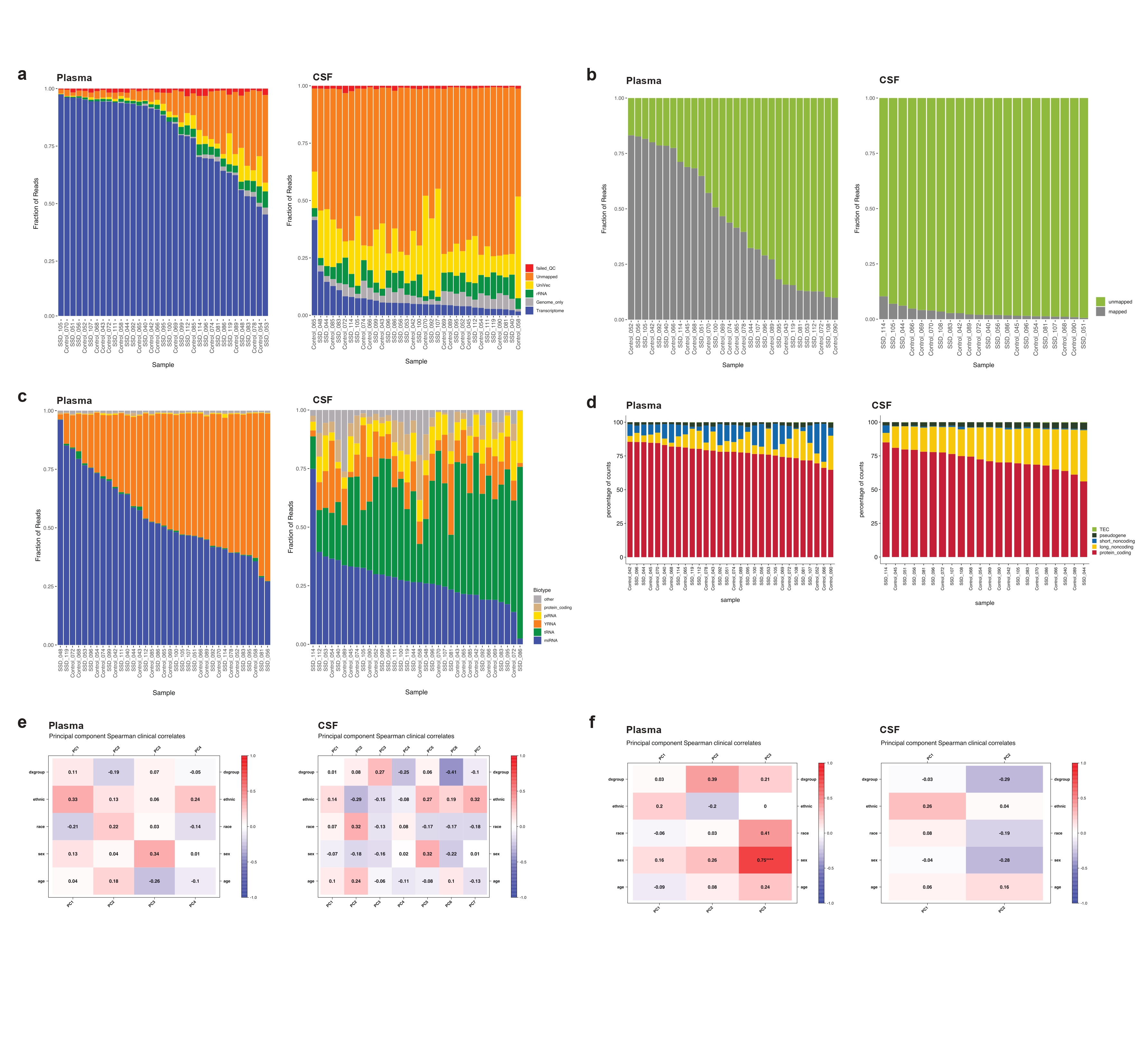

### Supplementary Figure 2.

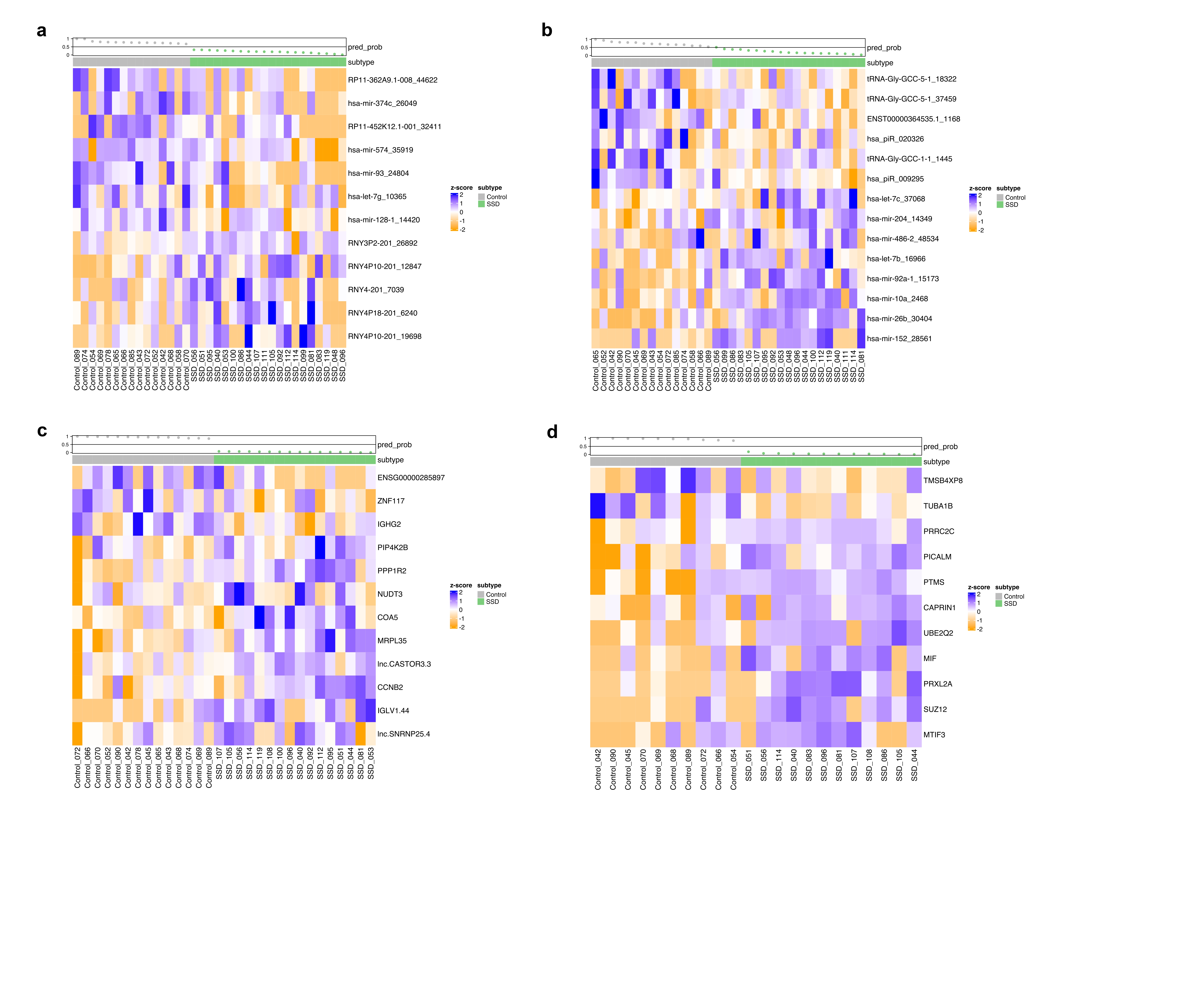
